# International risk of secondary hantavirus clusters following MV Hondius outbreak

**DOI:** 10.64898/2026.05.21.26353570

**Authors:** Boxuan Wang, Enrico Lorenzetti, Francesco Parino, Vittoria Colizza, Eugenio Valdano

## Abstract

The multinational Andes virus outbreak linked to the MV Hondius has exposed contacts across several countries, but the absence of further confirmed cases remains difficult to interpret given the long incubation period. We estimate the probability that secondary clusters may emerge using a stratified branching-process model parameterized with country-level tracing and isolation indicators. The risk of sustained spread is low, but secondary clusters remain plausible under imperfect isolation or pre-symptomatic transmission. These results support coordinated contact tracing and effective isolation while exposed contacts remain within the risk window.

## Body

An outbreak of Andes virus (ANDV) infection aboard the MV Hondius cruise ship has become the focus of international public-health attention. As of May 17, 10 cases had been reported, including eight confirmed and three deaths. ANDV can cause hantavirus pulmonary syndrome, a severe disease with high case fatality, and, unlike other hantaviruses, ANDV has documented capacity for person-to-person transmission, with estimates of the basic reproduction ratio R_0_ above one in previous outbreaks^1^. In response to the current outbreak, extensive tracing of contacts exposed before implementation of coordinated medical repatriation has been initiated, including passengers potentially exposed during international flights before confirmed cases were identified^2^. Isolation and follow-up practices currently differ across countries, ranging from hospital-based isolation to home monitoring alone. In parallel, uncertainty remains regarding the potential for pre-symptomatic transmission.

We adapted a stratified branching process model^3^ to estimate the probability of secondary transmission clusters (≥3 infections). We considered 30 countries with documented links to known cases, MV Hondius crew or passengers, or their contacts^2^,and parameterized national contact tracing and isolation capacity using the Global Health Security (GHS) Index^4,5^ *Early Detection and Reporting* score and *Facilities Capacity* score, respectively. Assuming that 10% transmissions occur before symptom onset and a conservative R_0_=1.5, the probability of secondary clusters ranged from 0.4% to 0.7%, depending on national contact tracing capacity, under fully effective isolation (Fig. 1A). Using higher R_0_ estimates from previous outbreaks^1^ increased this range to 0.8%-1.2% (Fig. 1B). When isolation was only 90% effective (Fig. 1C,D), probabilities rose to 1.5%-2.3% for lower R_0_ and 2.9%-4.4% for higher R_0_. Doubling the proportion of pre-symptomatic transmission further increased probabilities to as high as 3.1%-4.8% and 5.9%-9.6%, respectively. High effectiveness in isolation of symptomatic cases substantially reduced the probability of secondary cluster emergence. With no pre-symptomatic transmission, countries with isolation effectiveness of at least 80% kept the probability of secondary clusters below 4% in both R_0_ scenarios, irrespective of contact tracing capacity (Fig. 1E). Assuming 10% pre-symptomatic transmission instead required isolation effectiveness of at least 90% (Fig. 1F).

**Figure 1:**
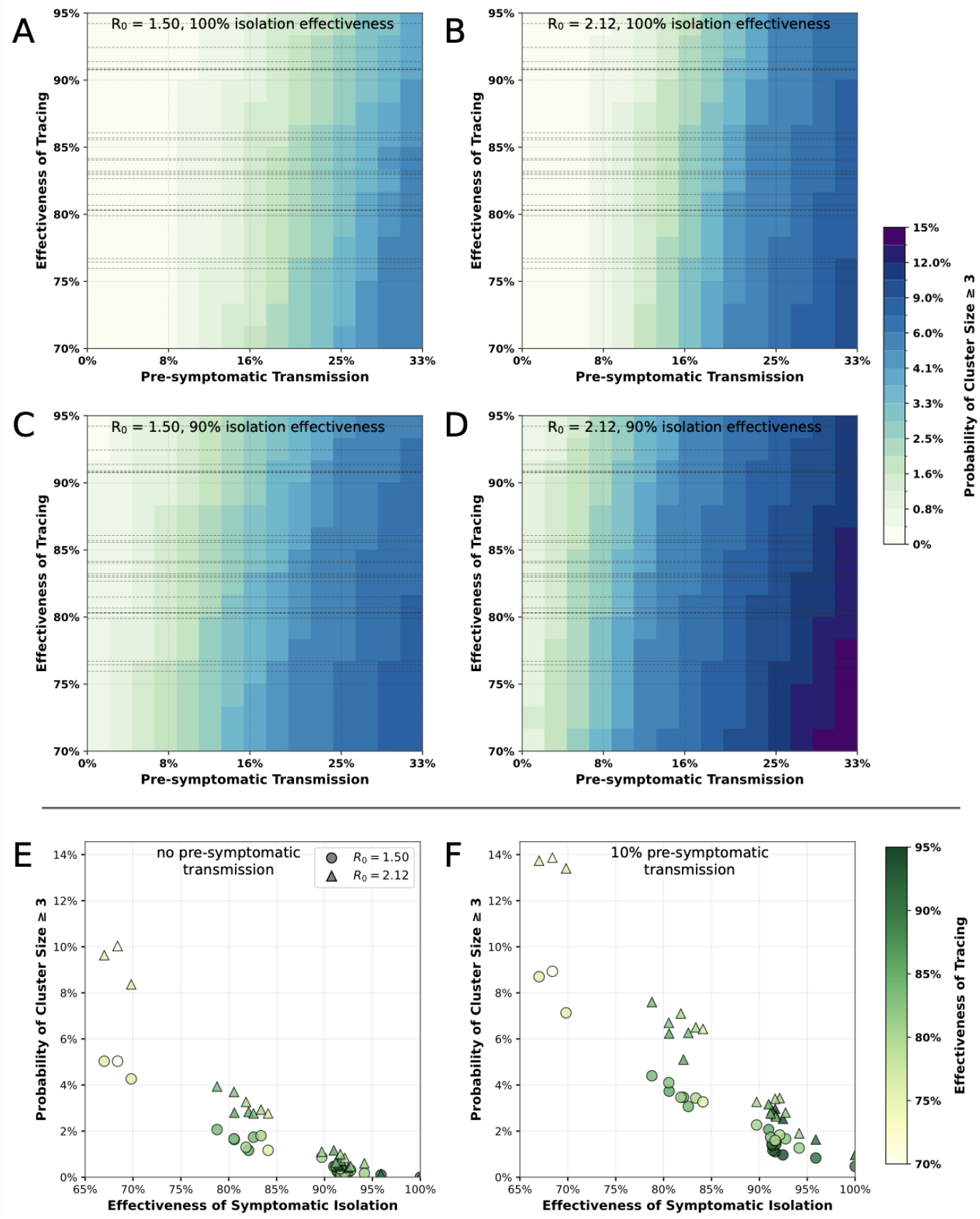
Risk of secondary clusters by pre-symptomatic transmission, tracing capacity and isolation effectiveness. A) Probability of cluster emergence as a function of the estimated pre-symptomatic transmission (fraction of transmission events occurring before symptom onset) and the effectiveness of tracing and isolation of contacts. 100% effectiveness means all contacts of a case are traced and isolated before they can infect. We used the Global Health Security (GHS) Index4 *Early Detection and Reporting* score as a proxy for each country’s test-and-trace capacity (countries indicated as horizontal lines). We set R_0_=1.5 and assumed that isolation of symptomatic cases is 100% effective in stopping all transmission since the onset of symptoms. B) Same as A but with R_0_=2.12 as estimated in Ref.^1^. C) Same as A but assuming that the isolation of symptomatic cases is 90% effective, i.e., prevents on average 9 in 10 transmission events occurring since the onset of symptoms. D) Same as B but assuming that the isolation of symptomatic cases is 90% effective. E) Probability of cluster emergence as a function of the effectiveness of isolation of symptomatic cases. We used The GHS index *Facilities Capacity* as a proxy for each country’s isolation capacity. The effectiveness of test-and-trace is also explored (color bar). R_0_=1.5 is shown in circles and R_0_=2.12 in triangles. We assumed no pre-symptomatic transmission. F) Same as E but assuming 10% pre-symptomatic transmission. Methodological description as well as supplementary results (alternative assumptions) are provided in the Appendix.

Although the probability of sustained transmission remains low, secondary clusters may still emerge under heterogeneous isolation practices and uncertain pre-symptomatic transmission. Maintaining rapid case identification, coordinated contact tracing, and effective isolation across countries may therefore be critical to limiting further international spread.

## Data Availability

All data produced are available online at https://ghsindex.org/.

## Acknowledgements

E.V. acknowledges the EU Horizon Europe grant SIESTA (101131957), the ANR JCJC grant DiscoReel (ANR-25-CE45-5346-01). V.C. acknowledges the EU Horizon Europe grant ESCAPE (101095619), the ANRS Maladies Infectieuses Émergentes and France 2030/SGPI grant PReViX (ANRS-24-PEPRMIE-0003). B.W., E.L., F.P., V.C., E.V. acknowledge the Working Group on Pandemic Preparedness of the French Research Action on Modeling Epidemics (FRAME - AC Modelisation) funded by the ANRS Maladies Infectieuses Émergentes.

