## Appendix for "International risk of secondary hantavirus clusters following MV Hondius outbreak"

#### Contents

**1 Materials and Methods** **2**

1.1 The transmission model . . . 2

1.2 Parametrization of contact tracing and isolation capacity . . . 2

**2 Supplementary results** **2**

### 1 Materials and Methods

#### 1.1 The transmission model

We model human-to-human transmission as a discrete-generation, stratified branching process. Strata are the combination of age classes ($[0, 18), [18, 40), [40, 60), [60, 80), [80, \infty)$) and the whether the contacts of a detected case are traced and isolated before they can transmit or not. The age-stratified contact matrix $C$ was sourced from Ref. [1] through `socialcontactdata.org`, and normalized so that its spectral radius matched the target $R_0$. We tested $R_0 = 2.12$ following Ref. [2] a lower value of $R_0 = 1.5$. We seeded the initial infection in the $[60, 80)$ age class, compatible with the cases of the MV Hondius. The number of secondary infections in age class $j$ of an infected individual in class $\ell$ that could occur in the absence of any control is sampled as follows:

$$X_{\ell j} \sim \text{NegBin}(\text{mean} = C_{\ell j}, \text{variance} = C_{\ell j}\,(1 + \omega C_{\ell j})). \tag{1}$$

We set $\omega = 0$ to model age-stratified community circulation. In Fig. 2 we tested a different value (see Sec. 2) Public-health control is represented as a thinning of this potential offspring distribution. Let $\phi$ be the probability that an infection is actually occurring, i.e., not prevented by control. This effectively models pre-symptomatic transmission, i.e., the probability that one infection is generated before symptom onset, in the case of prompt and effective isolation of symptomatic cases. It can also model infection after symptom onset but before effective isolation, i.e., isolation stopping all onward transmission. This gives

$$X_{\ell j}^{(\text{occurring})} \sim \text{Binomial}(X_{\ell j}, \phi). \tag{2}$$

The resulting infections are then split according to contact-tracing performance. There is a country-level parameter $p$ that encodes *contact tracing capacity*, i.e., the probability that, for each source individual, its secondary cases are traced and isolated before they can transmit. If they are, they will not generate additional infections, under the assumption that they are placed in effective pre-emptive isolation. Otherwise, they will contribute to onward transmission as described before.

#### 1.2 Parametrization of contact tracing and isolation capacity

We selected 30 countries that have relevant involvement with the occurring outbreak, either because their nationals were known to have been onboard the MV Hondius or been in contacts with cases or people who had been onboard the ship, or because visited by the ship, or because known to have had cases or contacts in their territories. They are Argentina, Australia, Belgium, Cabo Verde, Canada, Chile, France, Germany, Greece, Guatemala, India, Ireland, Italy, Japan, Montenegro, Netherlands, New Zealand, Philippines, Poland, Portugal, Russian Federation, South Africa, Spain, Switzerland, Taiwan, Turkey, Ukraine, United Kingdom, United States.

We used indicators from the Global Health Security (GHS) Index (`ghsindex.org`). We used the indicators *Early Detection and Reporting score* as a proxy for national capacity to trace contacts of cases and isolate them pre-emptively before they can transmit, to parametrize the parameter $p$.

We assumed a range $p_{min}$ and $p_{max}$ and then computed each country's $p$ from its indicator value $x$ through a linear interpolation:

$$p = p_{min} + \frac{p_{max} - p_{min}}{x_{max} - x_{min}} \left(x - x_{min}\right), \tag{3}$$

with $x_{max}, x_{min}$ computed over the selected countries. We selected $p_{min} = 0.7$ and $p_{max} = 0.95$.

As a proxy for countries capacity to enforce effective isolation of symptomatic cases we used the indicator *Facilities capacity*. We then assumed that no pre-symptomatic transmission occurred and parametrized $\phi$ through the indicators as the fraction of infection events occurring before effective isolation is in place. We used the same approach as before:

$$\phi = \phi_{min} + \frac{\phi_{max} - \phi_{min}}{y_{min} - y_{max}} \left(y_{min} - y\right), \tag{4}$$

with $y_{max}, y_{min}$ being this indicator computed over the selected countries. We selected $\phi_{min} = 0$ and $\phi_{max} = 0.33$.

### 2 Supplementary results

In Fig. 1 we report cluster emergence probabilities when setting a cutoff cluster size of 4 instead of 3. In Fig. 2 we test the effect of assuming the overdispersion reported in Ref. [2]. That value, however, however, was estimated in superspreading events and on age-unstratified data. In any case, the different choice had a minimal impact on the results.

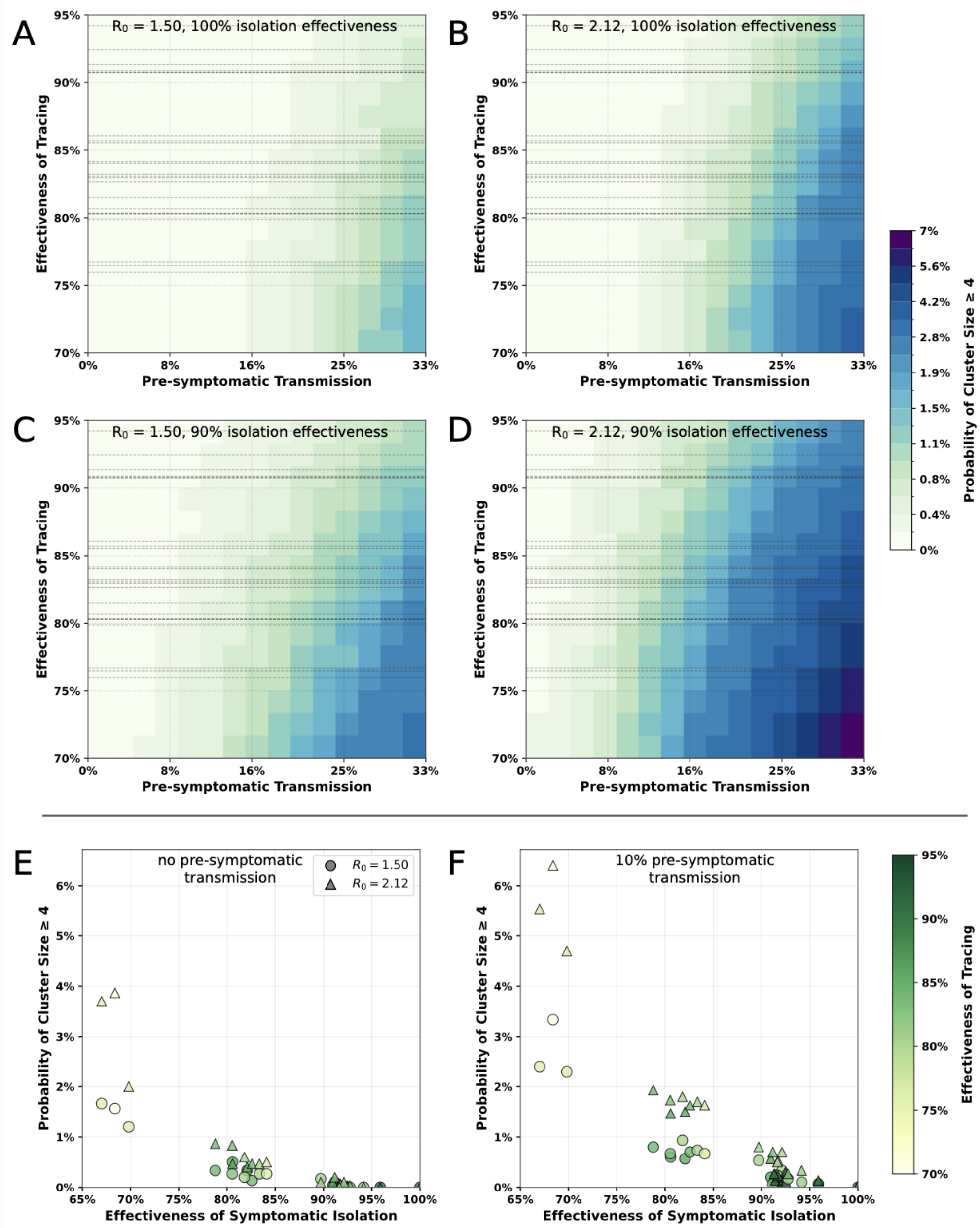


Figure 1: Same as the figure of the main text, but with cutoff cluster size set at ≥ 4.

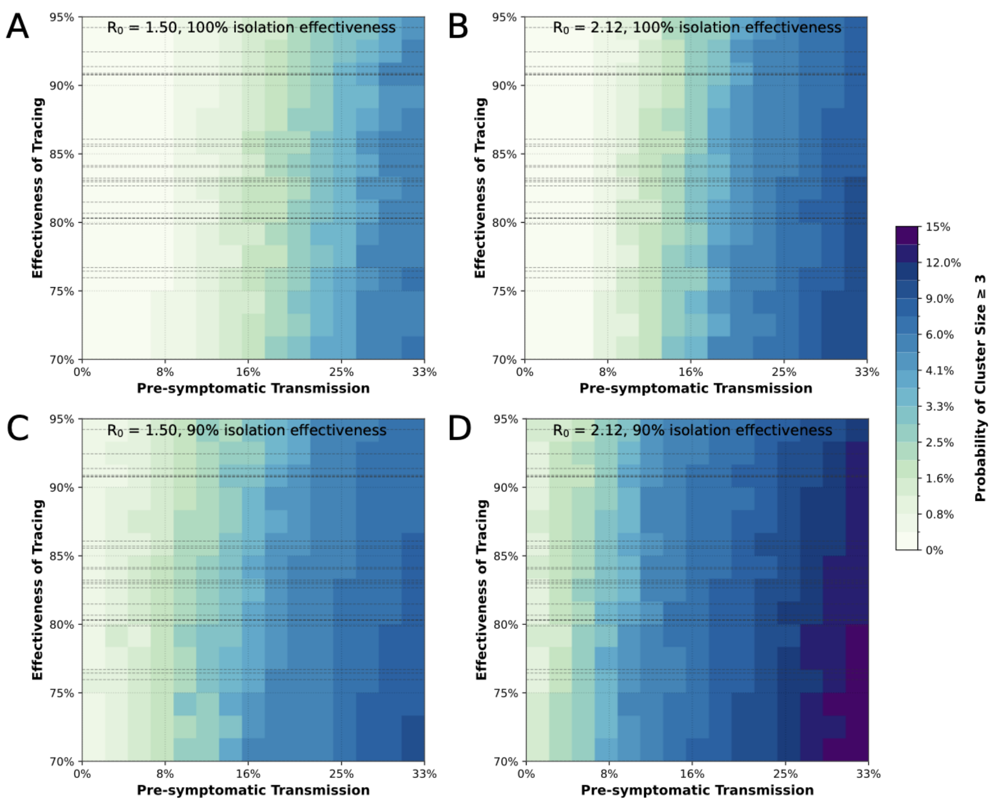


Figure 2: Same as the figure of the main text (panels A-D), but setting overdispersion parameter $\omega = 1.6$ (see Ref. [2]).
